# Validation of neutralizing antibody titers for estimating vaccine effectiveness for the Omicron SARS-CoV-2 variant, BA.1

**DOI:** 10.1101/2021.12.10.21267594

**Authors:** Billy J. Gardner, A. Marm Kilpatrick

## Abstract

**Background:** The emergence of new virus variants, including the Omicron variant (B.1.1.529) of SARS-CoV-2, can lead to immune escape and reduced vaccine effectiveness. Neutralizing antibody titers could be used to quickly estimate vaccine effectiveness (VE), because they can be easily measured following the emergence of a new virus variant and have been shown to be a correlate of protection for SARS-CoV-2 and other pathogens. However, few studies have examined VE-neutralizing antibody titer relationships with multiple virus variants, and none have validated relationships for immune evasive variants.

**Methods:** We leveraged variation among vaccines and virus variants to estimate VE-neutralizing antibody titer relationships across a 54-fold range of neutralizing antibody titers for two endpoints for COVID-19: symptomatic disease, and hospitalization. We predicted VEs for Omicron three days after the first neutralizing antibody titer became available. We tested these predictions using subsequently collected observational VE data.

**Findings:** For two mRNA vaccines (mRNA-1273, BNT162b2), fitted models predicted that infection with the BA.1 Omicron variant would increase the risk of hospitalization 2.8-4.4-fold and increase the risk of symptomatic disease 1.7-4.2-fold compared to the Delta variant. However, a third vaccine dose was predicted to restore protection. Out-of-sample validation data indicated that model predictions were quite accurate, with all predictions being within 10% of observed VE estimates, and all empirical estimates fell within the model prediction intervals.

**Interpretation:** These analyses demonstrate that models using neutralizing antibody titers can provide rapid VE estimates which can inform vaccine design and selection.

**Funding:** California Department of Health, National Science Foundation

## Introduction

The evolution of pathogens in populations with high immunity from infection or vaccination can lead to new variants with substantial immune escape ^1–4^. Determining the extent of immune evasion on the effectiveness of vaccines is critical for assessing the need for new and variant-specific vaccines and determining if additional non-pharmaceutical interventions are needed to limit spread ^5^. Traditional vaccine effectiveness (VE) studies can only be performed when there is significant transmission of the new variant in a partially vaccinated population and can be costly. Delays in implementing interventions or developing vaccines until traditional VE studies can be performed could result in rapid growth of a new variant, stress on healthcare systems and substantial preventable disease and death ^5^. Developing and validating faster methods for estimating VE is therefore critical.

One way to estimate VE against new pathogen variants is to use a surrogate of protection. Protection against infection and severe disease has been correlated with immune responses for several pathogens. Neutralizing antibody titers have been shown to correlate with protection against all infections for several arboviruses, and against symptomatic disease for some viruses that replicate in the mucosae ^6^, including influenza ^7^. For some endemic human coronaviruses, IgA in the serum and mucosa were associated with shorter duration of viral shedding and neutralizing antibody titers were correlated with protection against symptomatic disease ^8^. More recently, neutralizing antibody titers were strongly correlated with protection against symptomatic disease for COVID-19 against the original virus that emerged in 2020 ^9^. Establishing correlates of protection can be used for updating vaccines with much smaller immunogenicity-based trials than the randomized control trials needed to establish vaccine efficacy ^6,10^. However, it’s unclear whether neutralizing antibody titers, which are relatively easy to measure, are accurate correlates of protection for severe disease and hospitalization, which are often influenced by T-cells ^6,11^. It’s also unclear whether surrogates such as neutralizing antibody titers can be used to accurately estimate protection for novel pathogen variants.

The emergence and rapid spread of the Omicron variant (B.1.1.529) of SARS-CoV-2 in November of 2021 in South African populations raised the possibility that this new variant was highly immune evasive ^12^. However, the immune landscape at this time was highly variable globally. In South Africa, recent infection-based immunity was high but vaccination-based immunity was low. In contrast, in many developed countries, a large fraction of the population had been vaccinated but immunity had waned since second doses were administered ^12–15^. VE estimates were urgently needed to determine the most effective public health response to the spread of the Omicron variant, which could include third vaccine doses, new vaccines, non-pharmaceutical interventions, or new treatments. Waiting for enough cases to accumulate to estimate VE using observational studies could lead to rapid spread of the Omicron variant, stress on healthcare systems and substantial preventable disease and death. Previous studies had shown that variation in neutralizing antibody titers across vaccines was strongly correlated with protection against symptomatic disease, all infections, and transmission, but only for non-immune evasive virus variants ^9,14,16,17^. Here, we extend this approach by examining the accuracy of predicting VE for the highly immune evasive Omicron variant for both mild and severe disease, a key vaccine endpoint ^11^.

Our goal was to determine the accuracy of neutralizing antibody titers in predicting VE for mild and severe disease for the new Omicron virus variant. First, we examined variation in neutralizing antibody titers to determine the extent of data that is needed to accurately measure this surrogate of protection for a new virus variant. Second, we examined the relationships between neutralizing antibody titers and VE for two commonly measured and important endpoints, hospitalization and symptomatic disease, using data for 7 vaccines and 5 virus variants, including the immune evasive Beta variant. Third, we used the relationships to predict VE for the Omicron variant for these endpoints. Finally, we assessed the accuracy of these predictions by comparing model predictions with subsequently collected empirical VE estimates. We show that neutralizing antibody titers are an accurate surrogate of protection to quickly estimate VE for both mild and severe disease during a public health emergency.

## Methods

### Relative neutralizing antibody titers by variant, vaccine dose and with waning

We collected data on neutralizing antibody titers for SARS-CoV-2 variants, including Wild Type (WT) (including D614G), Alpha, Beta, Gamma, Delta, and three Omicron variant sub-lineages (BA.1, BA.2, and BA.4/5) (Table S1). For each vaccine in a study, we normalized neutralizing antibody titers for WT virus, T_vac,WT_, by dividing them by the neutralizing antibody titers of convalescent sera from that study T_conv,WT_ to produce a neutralizing antibody titer ratio, NATR_vac_ ^9^:

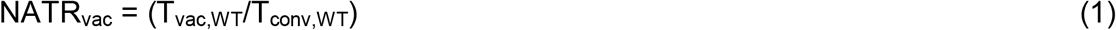

This produces a measure of the ability of a vaccinated person’s blood to neutralize the wild-type virus relative to blood of a person that has been infected with wild-type virus. For example, vaccination with BNT162b2, made by Pfizer-BioNTech, resulted in neutralizing antibody titers for wild-type (WT) SARS-CoV-2 that were approximately 2.37-fold higher than convalescent sera, producing a NATR_vac_ of 2.37 ^9^.For each non-WT variant, we divided the neutralizing antibody titer for a vaccine, T_vac,var_ for that variant by the neutralizing antibody titer for that vaccine for wild-type (WT) SARS-CoV-2 (including Wuhan-Hu-1, US-WA1/2020, B, and B.1 lineages), T_vac,WT_. This produced a ratio representing the reduction (often expressed as an X-fold difference) between a virus variant and WT virus, NATR_var_:

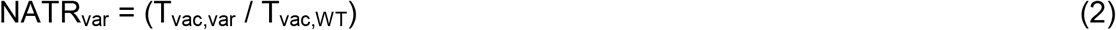

We examined neutralizing antibody titers and VE shortly after initial vaccination and for two additional immune statuses for mRNA vaccines: *waned* two dose projection (>6 months post 2^nd^ dose) and recently *boosted* (2-4 weeks) after a third vaccine dose. We used the relative neutralizing antibody titer ratio for these two immune statuses relative to recent two-dose neutralizing antibody titers to produce a third neutralizing antibody titer ratio:

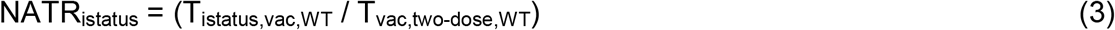

NATR_waned_ was similar for both mRNA vaccines: 0.124 (95% CI: 0.088-0.138) for BNT162b2 and 0.118 (95% CI: 0.115-0.120) for mRNA-1273, indicating that neutralizing antibody titers were approximately 8-fold lower 6+ months after vaccination. NATR_boosted_ was 1.54 (95% CI: 1.52-1.57) for mRNA-1273 and 3.22 (95% CI: 2.82-4.24) for BNT162b2 ^14^. The total relative neutralizing antibody titer ratio, NATR_tot_, for each vaccine, virus variant, and immune status is:

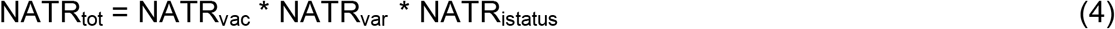

The first term in eq. 4 is the ratio previously used as surrogates of protection for symptomatic disease for COVID-19 from WT-virus ^9^ and the second term is the inverse of the “X-fold reduction” in neutralization of a non-WT variant relative to WT. The third term is the fold-change for a given immune status relative to titers immediately following a second vaccine dose.

In estimating NATR_var_, we only included studies that measured titers for WT virus and at least 2 other virus variants to reduce between-lab differences in titers for single variants (see below). We examined differences among virus variants using a linear mixed effects model with variant and vaccine as fixed effects and study ID as a random effect using the *lme4* package using *R, v4*.*2*.*2*.

### VE against symptomatic disease and hospitalization

We collected VE estimates for COVID-19 from the literature (including a systematic living review ^18^) and categorized each study by vaccine, variant type (Alpha, Beta, Gamma, Delta,), and endpoint (hospitalization and symptomatic disease). We excluded estimates where the virus variant for the VE estimate could not be determined.

To fit a relationship between VE and NATR, we needed estimates of the number of infections in the vaccine and control groups. However, most studies did not report the number of infections; however, all reported a VE and 95% CI. We estimated the effective number of infected individuals in the control group (I_c_) and the effective number of infected individuals in the vaccine group (I_v_) for each study by determining the number of each needed to match the mean and 95% CI given in a study. We held the effective number of individuals in the control group (N_c_) and vaccine group (N_v_) constant at 1,000,000 because the 95% CI was invariant to variation in these values for observed incidence values. We used a maximum value of 1000 infections in the control group (I_c_) to reduce the undue leverage of some very large studies ^19^.

### Relationships between VE and neutralizing antibody titers by vaccine and variant

We modeled the relationship between NATR_tot_ and VE for each endpoint across all vaccines and variants as:

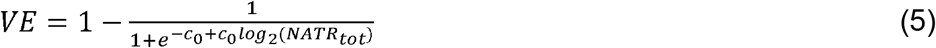

This mathematical form constrains the mean VE estimate to be between 0 and 1 because we believed it was unlikely that VE would be negative even for very low neutralizing antibody titers. We estimated c_0_ and c_1_ separately for the two endpoints, symptomatic disease and hospitalization, by maximizing the likelihood of observing the number of infections in the control group, I_c_, and vaccine group, I_v_, for each study, where the likelihood was a product of two binomial distributions,

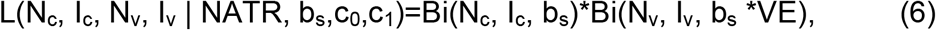

Here b_s_ is the baseline risk for the study period, which is the fraction of control individuals infected during the study. We used the fitted relationships between VE and neutralizing antibody titer to estimate VE for the Omicron subvariant BA.1 for the two endpoints, symptomatic disease and hospitalization.

We report two VE predictions for the BA.1 subvariant of Omicron using two values of NATR_var_. When we first posted a preprint on December 11, 2021, we used a 39 fold-reduction in neutralizing antibody titers (NATR_var_ = 1/39), based on the first unpublished data available on December 8, 2021 ^20,21^. Subsequent studies suggested a lower value (NATR_var_ = 1/18.5); we also show predictions using this value.

### Validating predictions

We collected data from the literature on VE estimates for the Omicron subvariant BA.1: for two vaccines (mRNA-1273 and BNT162b2), two immune statuses (for people recently vaccinated with a third dose and those with waned immunity, > 6 months after two doses), and two endpoints, symptomatic disease and hospitalization. We compared our VE predictions to the validation data; when there were multiple estimates for one of the eight possibilities, we calculated a weighted mean using the inverse of the variance as weights.

All code and data to replicate the results can be found at https://github.com/marmkilpatrick/New-Variant-VE.

## Results

There was substantial variation in the NATR_var_ measures for a single virus variant (5-10-fold), but relative variation in NATR_var_ measures across variants was much smaller (Fig. 1; Table S1). For example, while there was a 4-5-fold range in NATR_var_ measures for each variant, the rank order and relative differences in NATR_var_ measures between variants was quite consistent (Fig. 1). Of the variants we considered, Alpha was the least immune evasive, with only a 1.5-fold reduction in neutralizing antibody titers relative to WT (NATR_var_ = 0.64; 95% CI: 0.46-0.88) and Beta was the most immune evasive before the Omicron variant arose (NATR_var_ = 0.16; 95% CI: 0.12-0.22) (Fig. 1). Initial neutralizing antibody data for the Omicron variant (BA.1) from mid-December 2021 suggested an 39-fold reduction in neutralizing antibody titers (NATR_var_ = 0.026; 95% CI: 0.010-0.063) (Fig. 1 rightmost points), whereas subsequent data indicated lower immune evasion (18.5-fold or NATR_var_ = 0.054; 95% CI: 0.050-0.073) (Fig. 1).

**Fig. 1.**
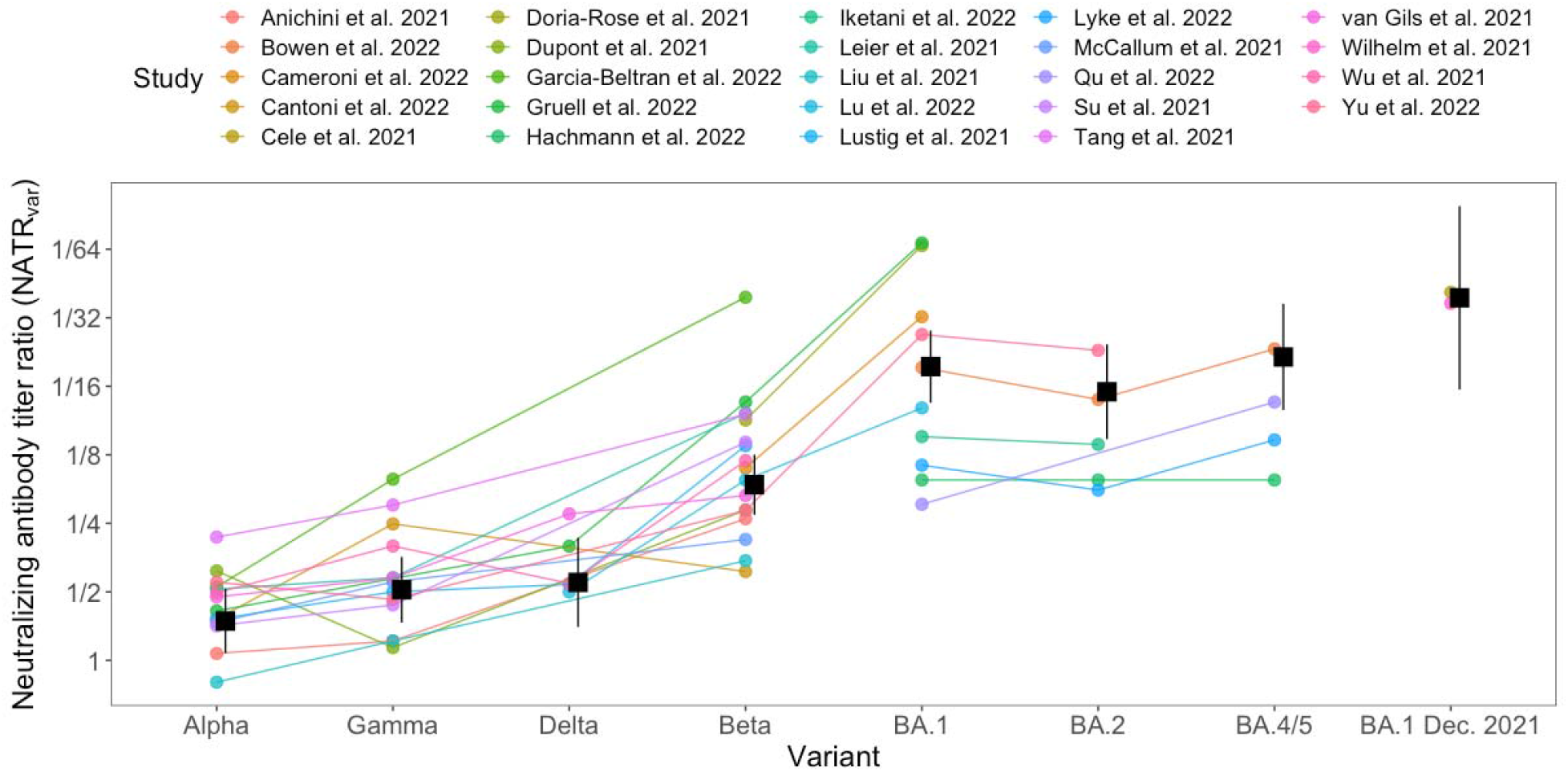
Neutralizing antibody titer ratios for virus variants relative to WT virus (NATR_var_) for five SARS-CoV-2 variants and three subvariants. Each point shows the reduction in neutralizing antibody titer relative to WT virus for a specific vaccine against a virus variant. Colors and lines connect points from the same study. Black squares and error bars show the mean and 95% confidence interval for each variant from the fitted model.

There were strong relationships between VE and NATR_tot_ for both symptomatic disease and hospitalization, which enabled us to estimate VE for new virus variants for multiple immune statuses, including waned immunity (Fig. 2; Tables S4, S5). These strong relationships were due, in part, to the 54- and 11-fold ranges in neutralizing antibody titers for symptomatic disease and hospitalization, respectively, that existed when we used data from all variants and vaccines together (Fig. 2). In contrast, for most individual vaccine-endpoint combinations, data was sparse and there were weak relationships between VE and NATR_tot_ (Figs. S1, S2).

**Fig. 2.**
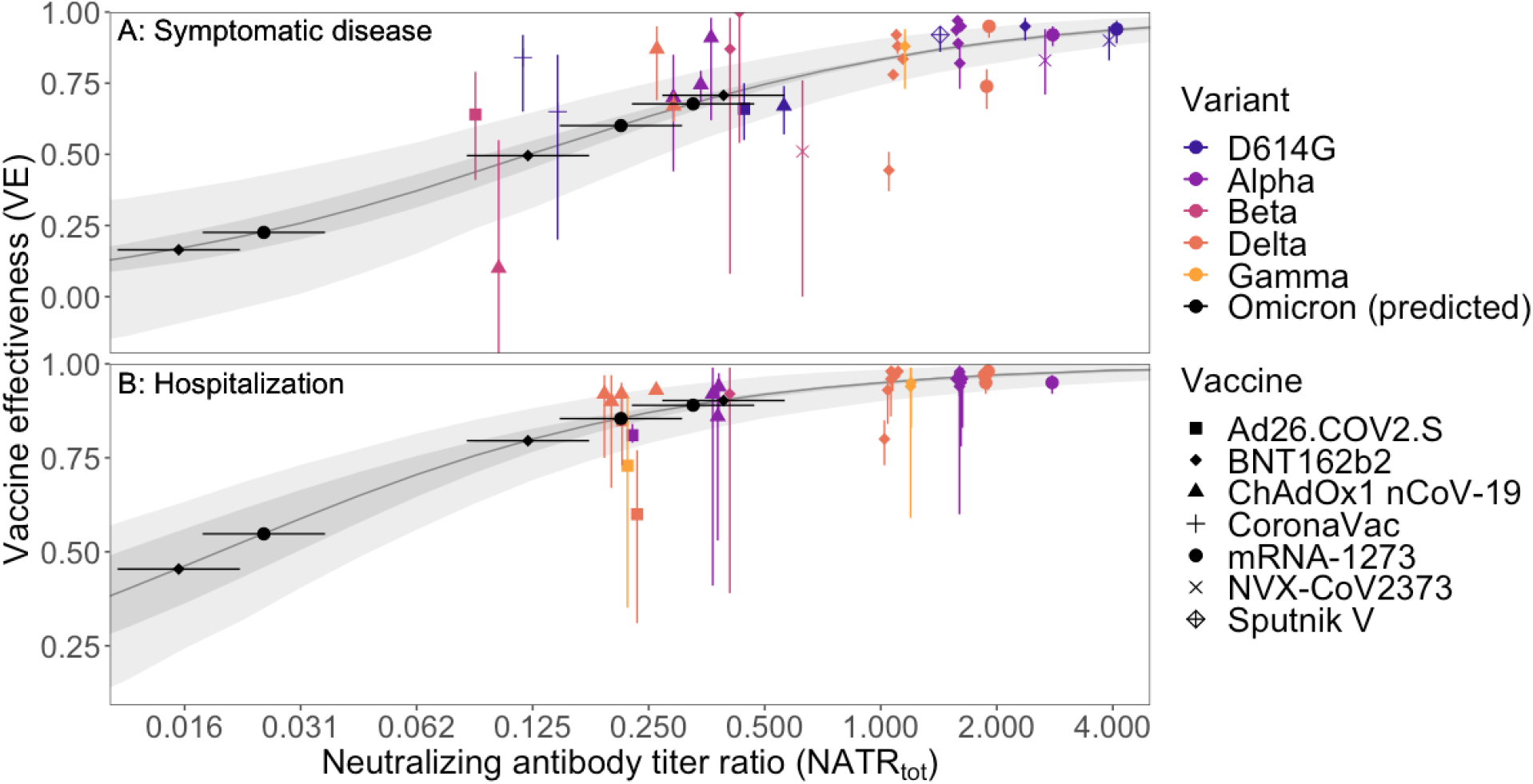
Vaccine Effectiveness (VE) plotted against variant- and vaccine-specific neutralizing antibody titer ratios (NATR_tot_) for (A) symptomatic disease and (B) hospitalization. Each point (and 95% CI), except those for Omicron predictions, represents a single empirical estimate of VE for a single vaccine & virus variant. Points are jittered slightly along the x-axis to facilitate presentation. Black points show predicted values for the Omicron variant using the fitted model; horizontal error bars show 95% CIs for Omicron-specific neutralizing antibody titer ratios. Dark ribbons show 95% CIs for the fitted model, and lighter ribbons show the 95% prediction intervals with 1000 infections in the unvaccinated control group.

We used the relationships between VE and NATR_tot_ (Figure 2) to predict VE for two mRNA vaccines, BNT162b2 and mRNA-1273, for the BA.1 subvariant of Omicron for waned (two-dose) immunity and after a third-dose booster (Figs. 2,3). We posted a preprint with these VE estimates on December 11, 2021 ^22^, just 3 days after the first neutralizing antibody titer ratio (NATR_var_) estimates for Omicron became available (December 8, 2021).

**Fig. 3.**
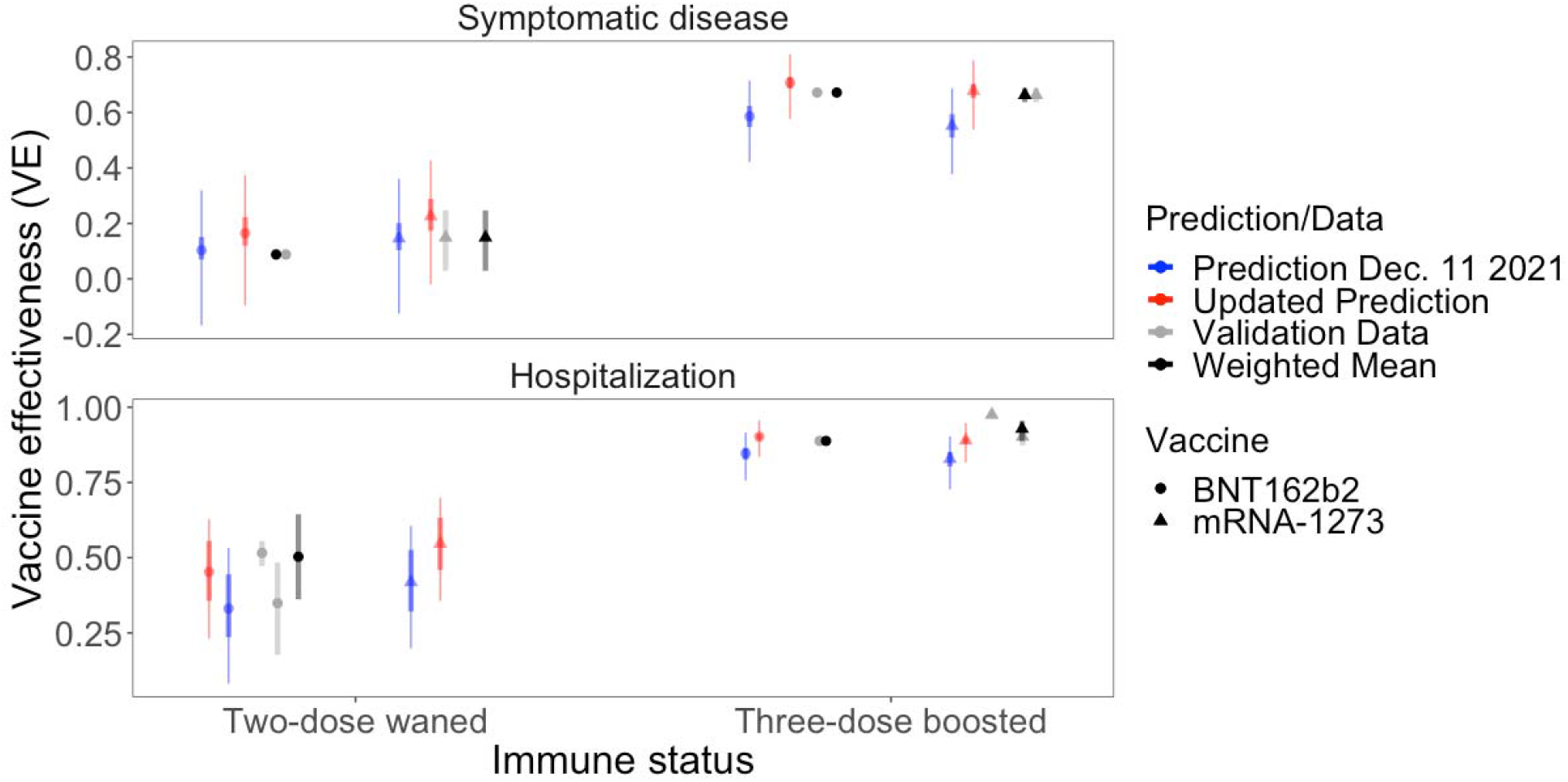
Predicted vaccine effectiveness (thick errors bars are 95% CIs; thinner error bars are 95% PIs), VE, for the Omicron variant (BA.1) and observed validation data (and 95% CI), including weighted means, for eight VEs: two endpoints, two vaccines, and two immunity statuses (two-dose waned) and (three-dose boosted)). We show VE predictions for both initial NATR_var_ estimates available Dec 8, 2021 (1/39) (red) and using subsequent data (NATR_var_ = 1/18.5) (green).

The fitted model predicted that VE against hospitalization for the Omicron variant would be much lower than for the Delta variant, with relative risk of hospitalization (1-VE) increasing 2.8-4.4-fold for BNT162b2 and mRNA-1273 for both waned immunity and after a third dose booster (Figs. 2,3; Table S6). For example, VE for waned immunity,, which comprised the majority of vaccinees in many developed countries in late 2021 ^12–15^, against hospitalization for BNT162b2 for the Delta variant was 80.6% (95% CI: 77.5-83.3) but the fitted model predicted that it would be only 46.5% (95% CI: 37.1-56.3) for the Omicron variant, increasing relative risk 2.75-fold (19.4% to 53.5%) (Figs. 2,3).

However, the fitted model predicted that a third dose would reverse the loss in protection and increase VE to 90.7% (95% CI: 90.0-91.5) against the Omicron variant (Figs. 2,3). The reductions in VE for Omicron and the benefits of a third dose booster were similar for the mRNA-1273 vaccine (Figs. 2,3).

The fitted model predicted that VE against symptomatic disease would also be much lower for the Omicron variant than for the Delta variant (Figs. 2,3). For individuals with waned immunity VE for the BNT162b2 vaccine for symptomatic disease against the Delta variant was 51.6% (95% CI: 46.9-56.4) but the fitted model predicted that protection would be nearly eliminated against the Omicron variant (VE 17.5%, 95% CI: 12.8-23.5) (Fig. S3). However, as with protection against hospitalization, the fitted model predicted that boosting with a third vaccine dose would restore VE for symptomatic disease (72.1% (95% CI: 70.2-74.1) for BNT162b2, with similar effects for mRNA-1273; Fig. S3).

We compared VE predictions using initial estimates of NATR_var_ (1/39) and updated estimates (NATR_var_ = 1/18.5) for the BA.1 subvariant of Omicron to empirical VE estimates from subsequent observational studies for populations with waned immunity and two weeks after a third dose (Fig. 3). Weighted means of the validation data fell within the 95% PIs for all 7 predictions and all absolute VE errors were less than 10%. VE predictions against symptomatic disease were slightly higher than observed data for the four estimates but no bias was clear in VE predictions against hospitalization. Mean absolute error was 3.0% (sd = 0.9%) for hospitalization and 6.4% (sd = 3.0%) for symptomatic disease. Mean absolute errors were similar for the two vaccines, BNT162b2 (4.9%; sd = 2.9%) and mRNA-1273 (5.1%; sd = 3.5%).

## Discussion

The emergence and rapid growth of the Omicron (B.1.1.529) variant of SARS-CoV-2 in South Africa, with numerous known and novel mutations in the spike protein, created an urgent need to estimate VE against this virus ^12^. We used the relationships between NATR_tot_ and VE (Fig. 2) and NATR_var_ for Omicron to estimate VE for both symptomatic disease and hospitalization on 11 December, 2021, three days after the first estimates of NATR_var_ were made available on Twitter ^14^. Here, we have validated this approach for symptomatic disease and, importantly, severe disease, by showing that these estimates were consistent with subsequent empirical estimates based on observational data. In the process, we also identified two key components needed for accurately predicting VE for new virus variants using neutralizing antibody titer ratios.

First, we needed multiple measurements of NATR_var_ that measured NATR_var_ for multiple virus variants (not just the novel variant and WT virus). We found that single estimates of NATR_var_ varied almost an order of magnitude, while relative NATR_var_ among virus variants were much more repeatable (Fig. 1). Second, to link VE and NATR_tot_, we needed to combine data from multiple vaccines and virus variants to have a sufficiently wide range of VE and NATR_tot_ to estimate VE for an immune evasive variant like Omicron, especially for waned immunity (Fig 2). Future studies could use VE and NATR_tot_ for individuals with different immunity statuses (waned, recently vaccinated or with different number of doses) to broaden the range of VE and NATR to strengthen the relationship between VE and NATR_tot_.

The strong correlations between VE for hospitalization and NATR_tot_ using blood serum might, initially, be surprising, given the importance of many different arms of the immune system, including T-cells, in protection against severe disease ^11^. However, serum antibody levels frequently correlate with nasal antibody levels (possibly due to transudation from the blood to nasal mucosa) and nasal antibody levels play a key role in protection against infection ^16,23^. If antibodies prevent a person from becoming infected, they are also protected against severe disease. It is likely that the correlation between VE against hospitalization and NATR_tot_ partly results from neutralizing antibodies protecting against infection.

Using neutralizing antibody titers to provide initial estimates of VE is much faster than traditional VE studies (e.g. test-negative designs), and can be done for populations where traditional VE studies are difficult. These initial VE estimates can be used for vaccine selection, public health planning, determining whether to implement non-pharmaceutical interventions, and other aspects of public policy. The high accuracy of VE estimates using neutralizing antibody titers assessed with validation data for the BA.1 Omicron variant of SARS-CoV-2 suggests that this approach might be useful for other emerging pathogens (e.g., novel influenza virus strains). A key challenge is obtaining the requisite data needed to build relationships between VE and a surrogate of protection (e.g., neutralizing antibody titer) across a wide enough range to encompass the novel pathogen phenotype and host population immune status.

However, given the continued evolution of SARS-CoV-2, influenza and many other viruses, and the increased rate of spillover of novel pathogens into human populations, this is an important area for further research.

## Data Availability

All data produced in the present work are contained in the manuscript

