## Supplemental material for "Validation of neutralizing antibody titers for estimating vaccine effectiveness for the Omicron SARS-CoV-2 variant, BA.1"

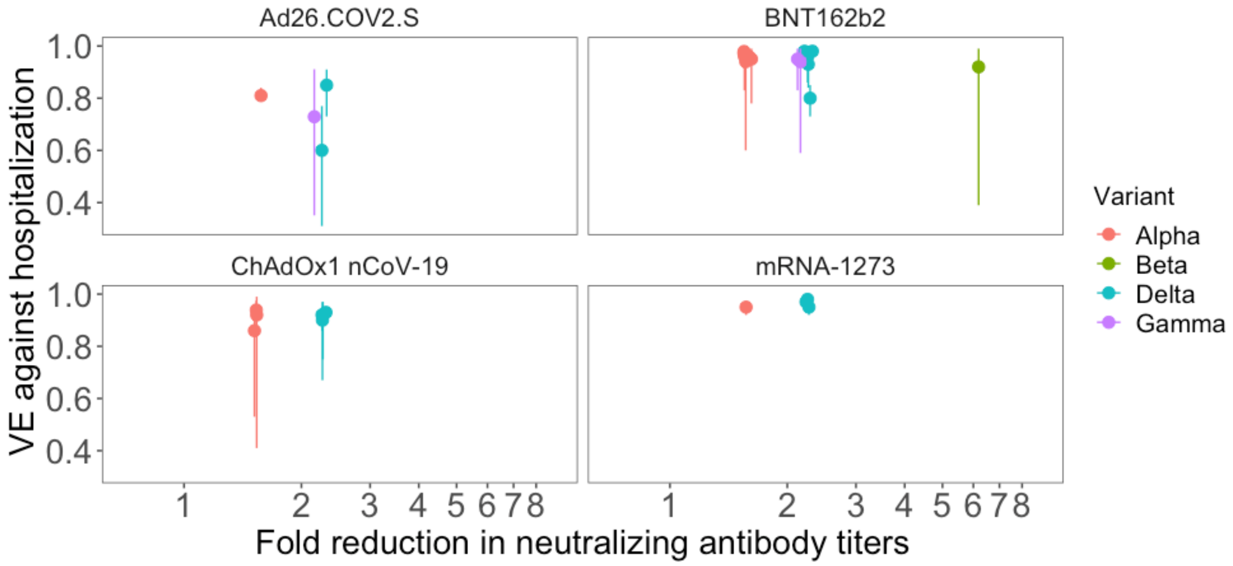


**Fig. S1. VE against hospitalization by vaccine plotted against variant-specific reductions in neutralizing antibody titers relative to WT virus. Each point (and 95% CI) represents a single estimate of VE for a single vaccine & virus variant from an observational study. Points have been slightly jittered along the x-axis to facilitate presentation.**

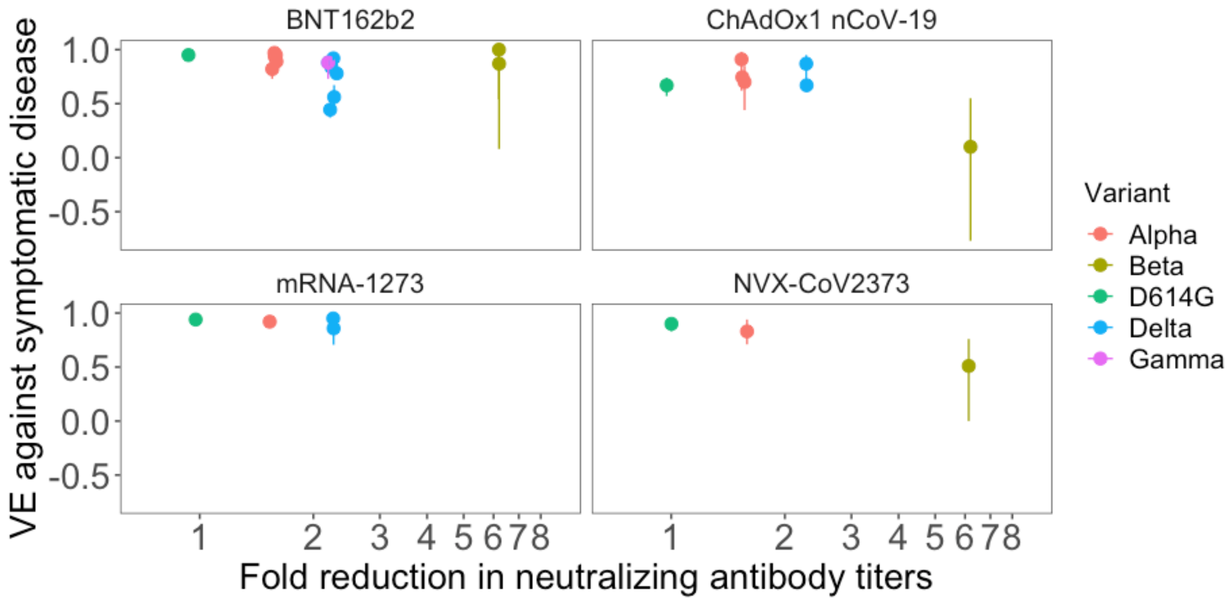


**Fig. S2. VE against symptomatic disease by vaccine plotted against variant-specific reductions in neutralizing antibody titers relative to WT virus. Each point (and 95% CI) represents a single estimate of VE for a single vaccine & virus variant from an observational study. Points have been slightly jittered along the x-axis to facilitate presentation.**

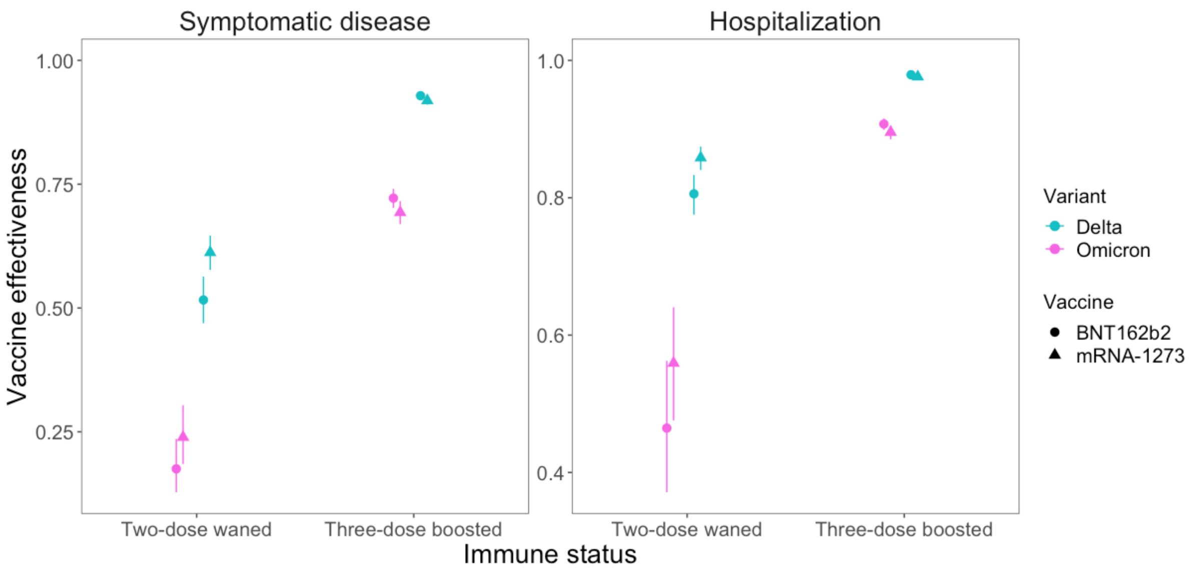


**Fig. S3. Comparison of estimated VE for the Omicron variant and Delta variants for two endpoints, two vaccines, and two immune statuses (6+ months after two doses (Waned) and shortly after a 3rd dose (Boosted)).**

**Table S1: Inverse of neutralizing antibody titers relative to WT (1/NATR$_{var}$).**

| Reference | Alpha | Gamma | Delta | Beta | BA.1 | BA.2 | BA.4/5 | BA.1 (Dec. 2021) |
|---|---|---|---|---|---|---|---|---|
| 1 | 1.1 | 1.2 | 0 | 4.2 | 0 | 0 | 0 | 0 |
| 2 | 0 | 0 | 0 | 0 | 19.3 | 14 | 23.3 | 0 |
| 3 | 0 | 0 | 0 | 7 | 32.3 | 0 | 0 | 0 |
| 4 | 1.5 | 4 | 0 | 2.5 | 0 | 0 | 0 | 0 |
| 5 | 0 | 0 | 0 | 0 | 0 | 0 | 0 | 41.4 |
| 6 | 0 | 0 | 7.7 | 0 | 0 | 0 | 0 | 0 |
| 7 | 0 | 0 | 0 | 11.4 | 66.3 | 0 | 0 | 0 |
| 8 | 2.5 | 1.1 | 0 | 4.6 | 0 | 0 | 0 | 0 |
| 9 | 2.1 | 6.3 | 0 | 39.4 | 0 | 0 | 0 | 0 |
| 10 | 1.6 | 0 | 3.2 | 13.7 | 68.3 | 0 | 0 | 0 |
| 11 | 0 | 0 | 0 | 0 | 6.2 | 6.2 | 6.2 | 0 |
| 12 | 0 | 0 | 0 | 0 | 9.6 | 8.9 | 0 | 0 |
| 13 | 2.1 | 2.3 | 0 | 12.2 | 0 | 0 | 0 | 0 |
| 14 | 0.8 | 1.2 | 0 | 2.7 | 0 | 0 | 0 | 0 |
| 15 | 0 | 0 | 2 | 6.2 | 12.9 | 0 | 0 | 0 |
| 16 | 1.5 | 2 | 2.2 | 8.8 | 0 | 0 | 0 | 0 |

| | | | | | | | | |
|---|---|---|---|---|---|---|---|---|
| 17 | 0 | 0 | 0 | 0 | 7.2 | 5.6 | 9.3 | 0 |
| 18 | 1.5 | 2.2 | 0 | 3.4 | 0 | 0 | 0 | 0 |
| 19 | 0 | 0 | 0 | 0 | 4.9 | 0 | 13.6 | 0 |
| 20 | 1.4 | 1.8 | 0 | 9.1 | 0 | 0 | 0 | 0 |
| 21 | 3.5 | 4.8 | 0 | 12.1 | 0 | 0 | 0 | 0 |
| 22 | 1.9 | 2.3 | 4.4 | 5.3 | 0 | 0 | 0 | 0 |
| 23 | 0 | 0 | 0 | 0 | 0 | 0 | 0 | 37 |
| 24 | 2 | 3.2 | 2.2 | 7.5 | 0 | 0 | 0 | 0 |
| 25 | 0 | 0 | 0 | 0 | 27 | 23 | 0 | 0 |
| 26 | 2.2 | 1.9 | 0 | 4.6 | 0 | 0 | 0 | 0 |

**Table S2: The fitted model for ln($NATR_{var}$) with variant a fixed effect and study as a random effect. Alpha was the reference level; the standard deviation of the random effect, study, was 0.51.**

| Predictor | Estimate | SE | t-value | P-value |
|---|---|---|---|---|
| Alpha (Intercept) | 0.45 | 0.16 | 2.75 | 0.0087 |
| Gamma | 0.32 | 0.16 | 2.05 | 0.048 |
| Delta | 0.37 | 0.23 | 1.63 | 0.11 |
| Beta | 1.37 | 0.15 | 9.26 | <0.001 |
| BA.1 | 2.47 | 0.22 | 11.06 | <0.001 |
| BA.2 | 2.17 | 0.29 | 7.49 | <0.001 |
| BA.4/5 | 2.54 | 0.31 | 8.25 | <0.001 |
| BA.1 Dec. 2021 | 3.22 | 0.49 | 6.63 | <0.001 |

**Table S3: Estimates of fold reduction in neutralizing antibody titers for virus variants relative to WT ($NATR_{var}$) with 95% CIs.**

| Variant | Fold Reduction (95% CI) |
|---|---|
| Alpha | 1.57 (1.14-2.16) |
| Gamma | 2.15 (1.55-2.99) |
| Delta | 2.27 (1.45-3.55) |
| Beta | 6.17 (4.58-8.30) |
| BA.1 | 18.5 (12.8-26.7) |
| BA.2 | 13.6 (8.3-22.4) |
| BA.4/5 | 19.9 (11.6-34.1) |
| BA.1 (Dec. 2021) | 39.1 (16.0-96.0) |

**Table S4: Vaccine effectiveness against hospitalization estimates used to fit the VE-NATR$_{tot}$ model and shown in Figure 2.**

| Variant | Reference | Vaccine | VE | 95% CI | Effective cases vaccine group | Effective cases control group |
|---|---|---|---|---|---|---|
| Alpha | [27] | Ad26.COV2.S | 0.81 | 0.79-0.84 | 190 | 1000 |
| Alpha | [28] | BNT162b2 | 0.96 | 0.83-0.99 | 1.9 | 46.9 |
| Alpha | [29] | BNT162b2 | 0.979 | 0.91-1 | 2 | 94 |
| Alpha | [29] | ChAdOx1 nCoV-19 | 0.939 | 0.85-0.98 | 5 | 81.6 |
| Alpha | [30] | BNT162b2 | 0.95 | 0.78-0.99 | 1.8 | 36.6 |
| Alpha | [30] | ChAdOx1 nCoV-19 | 0.86 | 0.53-0.96 | 3 | 21.3 |
| Alpha | [31] | BNT162b2 | 0.972 | 0.97-0.98 | 28 | 1000 |
| Alpha | [32] | BNT162b2 | 0.94 | 0.6-0.99 | 1.1 | 18.6 |
| Alpha | [33] | BNT162b2 | 0.96 | 0.94-0.97 | 27.3 | 681.3 |
| Alpha | [33] | mRNA-1273 | 0.95 | 0.92-0.97 | 17.8 | 356.3 |
| Alpha | [33] | ChAdOx1 nCoV-19 | 0.92 | 0.41-0.99 | 1 | 12.9 |
| Beta | [33] | BNT162b2 | 0.92 | 0.39-0.99 | 1 | 12.4 |
| Delta | [27] | Ad26.COV2.S | 0.85 | 0.73-0.91 | 12.9 | 85.8 |
| Delta | [34] | Ad26.COV2.S | 0.6 | 0.31-0.77 | 17.8 | 44.5 |
| Delta | [34] | BNT162b2 | 0.8 | 0.73-0.85 | 52.2 | 261 |
| Delta | [34] | mRNA-1273 | 0.95 | 0.92-0.97 | 17.4 | 347.5 |
| Delta | [28] | BNT162b2 | 0.98 | 0.97-0.98 | 20 | 1000 |

| | | | | | | |
|---|---|---|---|---|---|---|
| Delta | [28] | mRNA-1273 | 0.97 | 0.96-0.98 | 30 | 1000 |
| Delta | [28] | ChAdOx1 nCoV-19 | 0.92 | 0.86-0.95 | 13.5 | 169.2 |
| Delta | [29] | BNT162b2 | 0.967 | 0.96-0.97 | 33 | 1000 |
| Delta | [29] | ChAdOx1 nCoV-19 | 0.93 | 0.92-0.94 | 70 | 1000 |
| Delta | [35] | BNT162b2 | 0.93 | 0.84-0.96 | 6 | 85.8 |
| Delta | [36] | BNT162b2 | 0.976 | 0.93-0.99 | 3.3 | 135.6 |
| Delta | [30] | BNT162b2 | 0.96 | 0.86-0.99 | 2.6 | 65 |
| Delta | [30] | ChAdOx1 nCoV-19 | 0.92 | 0.75-0.97 | 3 | 37.7 |
| Delta | [33] | BNT162b2 | 0.98 | 0.96-0.99 | 8.2 | 410.4 |
| Delta | [33] | mRNA-1273 | 0.98 | 0.93-1 | 2.5 | 123.6 |
| Delta | [33] | ChAdOx1 nCoV-19 | 0.9 | 0.67-0.97 | 2.9 | 28.7 |
| Gamma | [28] | BNT162b2 | 0.95 | 0.83-0.99 | 2.7 | 53.6 |
| Gamma | [37] | Ad26.COV2.S | 0.729 | 0.35-0.91 | 6.2 | 23 |
| Gamma | [33] | BNT162b2 | 0.94 | 0.59-0.99 | 1.1 | 18.3 |

**Table S5: Vaccine effectiveness against symptomatic disease estimates used to fit the VE-NATR$_{tot}$ model and shown in Figure 2.**

| Variant | Reference | Vaccine | VE | 95% CI | Effective cases vaccine group | Effective cases control group |
|---|---|---|---|---|---|---|
| Alpha | [31] | BNT162b2 | 0.97 | 0.97-0.97 | 30 | 1000 |
| Alpha | [32] | BNT162b2 | 0.82 | 0.73-0.88 | 27.9 | 155.2 |
| Alpha | [33] | BNT162b2 | 0.89 | 0.87-0.9 | 110 | 1000 |

| | | | | | | | |
|---|---|---|---|---|---|---|---|
| Alpha | [33] | mRNA-1273 | 0.92 | 0.88-0.95 | | 24.7 | 308.2 |
| Alpha | [33] | ChAdOx1 nCoV-19 | 0.91 | 0.62-0.98 | | 2 | 22.4 |
| Alpha | [38] | BNT162b2 | 0.937 | 0.92-0.95 | | 49.7 | 788.2 |
| Alpha | [29] | BNT162b2 | 0.95 | 0.94-0.96 | | 50 | 1000 |
| Alpha | [38] | ChAdOx1 nCoV-19 | 0.745 | 0.68-0.79 | | 104.5 | 409.7 |
| Alpha | [39] | NVX-CoV2373 | 0.83 | 0.71-0.94 | | 14.1 | 82.7 |
| Alpha | [40] | ChAdOx1 nCoV-19 | 0.7 | 0.44-0.85 | | 12.6 | 42.1 |
| Beta | [33] | BNT162b2 | 0.87 | 0.08-0.98 | | 1.1 | 8.7 |
| Beta | [33] | BNT162b2 | 1 | 0.54-1 | | 0 | 17.8 |
| Beta | [41] | Ad26.COV2.S | 0.64 | 0.41-0.79 | | 21 | 58.2 |
| Beta | [42] | NVX-CoV2373 | 0.51 | 0-0.76 | | 11.2 | 22.9 |
| Beta | [43] | ChAdOx1 nCoV-19 | 0.1 | -0.77-0.55 | | 15.7 | 17.5 |
| D614G | [41] | Ad26.COV2.S | 0.66 | 0.55-0.75 | | 62.2 | 182.9 |
| D614G | [44] | ChAdOx1 nCoV-19 | 0.67 | 0.57-0.74 | | 81.9 | 248.1 |
| D614G | [45] | CoronaVac | 0.84 | 0.65-0.92 | | 7.5 | 47 |
| D614G | [46] | CoronaVac | 0.65 | 0.20-0.85 | | 7.4 | 21.2 |
| D614G | [47] | mRNA-1273 | 0.94 | 0.89-0.97 | | 10.7 | 177.9 |
| D614G | [48] | NVX-CoV2373 | 0.9 | 0.83-0.95 | | 14.5 | 145.4 |
| D614G | [49] | BNT162b2 | 0.95 | 0.90-0.98 | | 8.2 | 164.2 |
| D614G | [50] | Sputnik V | 0.92 | 0.86-0.95 | | 13.9 | 173.5 |
| Delta | [33] | BNT162b2 | 0.92 | 0.90-0.94 | | 69.6 | 869.9 |

| | | | | | | |
|---|---|---|---|---|---|---|
| Delta | 33 | mRNA-1273 | 0.95 | 0.91-0.97 | 11.6 | 233 |
| Delta | 33 | ChAdOx1 nCoV-19 | 0.87 | 0.69-0.95 | 5.8 | 44.5 |
| Delta | 38 | BNT162b2 | 0.88 | 0.85-0.90 | 108.3 | 902.5 |
| Delta | 51 | BNT162b2 | 0.444 | 0.37-0.51 | 381 | 685 |
| Delta | 29 | BNT162b2 | 0.835 | 0.83-0.84 | 165 | 1000 |
| Delta | 52 | BNT162b2 | 0.78 | 0.78-0.79 | 220 | 1000 |
| Delta | 51 | mRNA-1273 | 0.739 | 0.66-0.80 | 67.0 | 257 |
| Delta | 38 | ChAdOx1 nCoV-19 | 0.67 | 0.61-0.72 | 201.2 | 609.8 |
| Gamma | 33 | BNT162b2 | 0.88 | 0.73-0.94 | 6.4 | 53.3 |

**Table S6: Predicted VE, 95% CIs and 95% PIs for two vaccines, two variants, two endpoints, and two immune statuses using the model shown in Figure 2.**

| Variant | Prediction | Vaccine | Endpoint | Status | VE | 95% PI | 95% CI |
|---|---|---|---|---|---|---|---|
| Omicron | Updated | mRNA-1273 | Hospitalization | Three-dose boosted | 0.9 | 0.82-0.95 | 0.89-0.91 |
| Omicron | Updated | BNT162b2 | Hospitalization | Three-dose boosted | 0.91 | 0.84-0.96 | 0.9-0.92 |
| Omicron | Updated | mRNA-1273 | Hospitalization | Two-dose waned | 0.56 | 0.37-0.71 | 0.48-0.64 |
| Omicron | Updated | BNT162b2 | Hospitalization | Two-dose waned | 0.46 | 0.25-0.63 | 0.37-0.56 |
| Omicron | Updated | mRNA-1273 | Symptomatic disease | Three-dose boosted | 0.69 | 0.56-0.8 | 0.67-0.72 |
| Omicron | Updated | BNT162b2 | Symptomatic disease | Three-dose boosted | 0.72 | 0.6-0.82 | 0.7-0.74 |
| Omicron | Updated | mRNA-1273 | Symptomatic disease | Two-dose waned | 0.24 | -0.01-0.43 | 0.18-0.3 |
| Omicron | Updated | BNT162b2 | Symptomatic disease | Two-dose waned | 0.18 | -0.09-0.38 | 0.13-0.24 |

| | | | | | | | |
|---|---|---|---|---|---|---|---|
| Omicron | Updated | mRNA-1273 | All infections | Three-dose boosted | 0.69 | 0.56-0.8 | 0.66-0.72 |
| Omicron | Updated | BNT162b2 | All infections | Three-dose boosted | 0.71 | 0.57-0.81 | 0.68-0.73 |
| Omicron | Updated | mRNA-1273 | All infections | Two-dose waned | 0.44 | 0.21-0.62 | 0.33-0.55 |
| Omicron | Updated | BNT162b2 | All infections | Two-dose waned | 0.38 | 0.14-0.59 | 0.27-0.52 |
| Omicron | Dec. 11 2021 | mRNA-1273 | Hospitalization | Three-dose boosted | 0.83 | 0.73-0.91 | 0.81-0.85 |
| Omicron | Dec. 11 2021 | BNT162b2 | Hospitalization | Three-dose boosted | 0.85 | 0.76-0.92 | 0.83-0.87 |
| Omicron | Dec. 11 2021 | mRNA-1273 | Hospitalization | Two-dose waned | 0.42 | 0.19-0.6 | 0.33-0.52 |
| Omicron | Dec. 11 2021 | BNT162b2 | Hospitalization | Two-dose waned | 0.33 | 0.08-0.53 | 0.24-0.44 |
| Omicron | Dec. 11 2021 | mRNA-1273 | Symptomatic disease | Three-dose boosted | 0.56 | 0.39-0.7 | 0.52-0.6 |
| Omicron | Dec. 11 2021 | BNT162b2 | Symptomatic disease | Three-dose boosted | 0.59 | 0.43-0.72 | 0.56-0.63 |
| Omicron | Dec. 11 2021 | mRNA-1273 | Symptomatic disease | Two-dose waned | 0.15 | -0.11-0.36 | 0.11-0.21 |
| Omicron | Dec. 11 2021 | BNT162b2 | Symptomatic disease | Two-dose waned | 0.11 | -0.17-0.32 | 0.07-0.16 |
| Omicron | Dec. 11 2021 | mRNA-1273 | All infections | Three-dose boosted | 0.62 | 0.46-0.75 | 0.57-0.67 |
| Omicron | Dec. 11 2021 | BNT162b2 | All infections | Three-dose boosted | 0.64 | 0.49-0.76 | 0.6-0.68 |
| Omicron | Dec. 11 2021 | mRNA-1273 | All infections | Two-dose waned | 0.36 | 0.12-0.57 | 0.24-0.51 |
| Omicron | Dec. 11 2021 | BNT162b2 | All infections | Two-dose waned | 0.31 | 0.06-0.54 | 0.2-0.47 |
| Delta | Delta | mRNA-1273 | Hospitalization | Three-dose boosted | 0.98 | 0.94-1 | 0.97-0.98 |
| Delta | Delta | BNT162b2 | Hospitalization | Three-dose boosted | 0.98 | 0.95-1 | 0.97-0.98 |
| Delta | Delta | mRNA-1273 | Hospitalization | Two-dose waned | 0.86 | 0.77-0.93 | 0.84-0.87 |
| Delta | Delta | BNT162b2 | Hospitalization | Two-dose waned | 0.81 | 0.7-0.89 | 0.78-0.83 |

| | | | | | | | |
|---|---|---|---|---|---|---|---|
| Delta | Delta | mRNA-1273 | Symptomatic disease | Three-dose boosted | 0.92 | 0.86-0.97 | 0.91-0.93 |
| Delta | Delta | BNT162b2 | Symptomatic disease | Three-dose boosted | 0.93 | 0.87-0.97 | 0.92-0.94 |
| Delta | Delta | mRNA-1273 | Symptomatic disease | Two-dose waned | 0.61 | 0.45-0.74 | 0.58-0.65 |
| Delta | Delta | BNT162b2 | Symptomatic disease | Two-dose waned | 0.52 | 0.33-0.66 | 0.47-0.56 |
| Delta | Delta | mRNA-1273 | All infections | Three-dose boosted | 0.84 | 0.74-0.92 | 0.8-0.88 |
| Delta | Delta | BNT162b2 | All infections | Three-dose boosted | 0.85 | 0.75-0.93 | 0.81-0.89 |
| Delta | Delta | mRNA-1273 | All infections | Two-dose waned | 0.65 | 0.5-0.77 | 0.61-0.69 |
| Delta | Delta | BNT162b2 | All infections | Two-dose waned | 0.6 | 0.43-0.73 | 0.54-0.66 |

**Table S7: VE validation data for the Omicron variant shown in Figure 3 for two vaccines, two endpoints, and two immune statuses (two-dose waned and boosted with a third dose).**

| Reference | Vaccine | Dose | Endpoint | VE | CI | Estimated cases vaccine group | Estimated cases control group |
|---|---|---|---|---|---|---|---|
| [53] | mRNA-1273 | Three-dose boosted | Hospitalization | 97.5 | 96.3-98.3 | 25.9 | 1037 |
| [54] | BNT162b2 | Two-dose waned | Hospitalization | 51.6 | 47.2-55.6 | 776.8 | 1604.9 |
| [54] | BNT162b2 | Three-dose boosted | Hospitalization | 88.8 | 87.3-90.1 | 273.1 | 2438.4 |
| [54] | mRNA-1273 | Three-dose boosted | Hospitalization | 90.2 | 87.3-92.5 | 60.7 | 618.9 |
| [55] | BNT162b2 | Two-dose waned | Hospitalization | 34.9 | 17.7-48.4 | 116.2 | 178.5 |

| | | | | | | | |
|---|---|---|---|---|---|---|---|
| 56 | BNT162b2 | Two-dose waned | Symptomatic disease | 8.8 | 7 - 10.5 | 20033.7 | 21966.8 |
| 56 | BNT162b2 | Three-dose boosted | Symptomatic disease | 67.2 | 66.5 - 67.8 | 12151 | 37045.9 |
| 56 | mRNA-1273 | Two-dose waned | Symptomatic disease | 14.9 | 2.9 - 24.7 | 426.3 | 500.9 |
| 56 | mRNA-1273 | Three-dose boosted | Symptomatic disease | 66.3 | 63.7 - 68.8 | 902.5 | 2678 |
